# Source Matters: An Examination of Drug Checking Samples from Police Departments and Community Based Programs in Massachusetts

**DOI:** 10.64898/2026.05.08.26352755

**Authors:** Joseph Silcox, Sabrina S. Rapisarda, Ella Chase, Nick Huntington, Shannon Raeke, Amanda Consigli, Brandon del Pozo, Traci C. Green

## Abstract

**Aims and Setting:** In the U.S., the emergence of new adulterants and novel psychoactive substances continues to complicate approaches to overdose, treatment, and public safety. Information about this changing drug supply is often gleaned from police drug seizures, but community drug checking services, which test the contents of a person’s drug supply and share that data, provide another means to understand local drug supplies. However, it is unclear how seized drugs differ from those collected in the community, whether one approach is potentially more instructive, and what can be learned about local drug supplies from each source. We therefore compared drug samples tested from police departments (PDs) and community partner (CP) drug checking programs to examine what, if any, differences existed in sample content, form, submitter characteristics, and emerging substance presence.

**Design:** We conducted a retrospective cohort analysis of drug samples collected and tested between April 2018 and December 2025 by the Massachusetts Drug Supply DataStream derived from CPs and PDs operating in the same geographic area across eight locations. Bivariate analyses (Chi-square, Fisher’s exact) tested for differences in sample and submitter characteristics by source.

**Findings:** There were 2,430 unique samples submitted by CPs (68.1%) and PDs (31.9%) from the same location. Compared to CP samples, proportionally more PD samples showed fentanyl as primary substance (74.2% PD vs. 64% CP, p<.001) and less often contained additives (xylazine 15.0% PD vs. 27.4% CP; medetomidine 0.6% PD vs. 2.2% CP, both p<.001). PD samples were typically powders (73.2% vs. 37.9%) and pills (13.6% vs. 3.6%) while CP samples were more often residue (51.9% vs. 2.1%, p<.001). Submitter characteristics, when reported, differed by source: gender (n=528, male: 78.6% PD vs. 50.1% CP, p<.001), race/ethnicity (n=468, Black: 15.8% PD vs. 7.8% CP; Hispanic: 6.7% PD vs. 13.2% CP, p<.05), and associated overdose (n=242, fatal: 62.9% vs. 10.9%, p<.001). Emergent substances were detected a median of 249 days sooner in CP than co-located PD samples, though drugs exhibiting concerning patterns (e.g., unexpected fentanyl in stimulants) had similar, swift detection times.

**Conclusion:** Drug samples differ based on PD vs. CP source in significant ways that may introduce bias when drawing conclusions about drug supply trends but also offer unique insights for public health and responses to emerging drugs. Modern drug monitoring should include a broad range of sources to best prepare for changes the illicit supply may bring to overdose prevention, public safety, and health systems.

## INTRODUCTION

The overdose crisis persists as the leading cause of accidental death in the U.S., claiming over 400,000 lives in the past two decades (1–4). More recently, contaminants to the illicit opioid supply such as xylazine, medetomidine, and nitazenes have heightened overdose risk (5–9) by introducing complexity and unpredictability, and altering the signs, symptoms and effects of overdose, often without awareness among people who use drugs (PWUD) about the adulteration of the substances they consume (8,10).

A major barrier to overdose prevention is the lack of timely data about a local drug supply. Knowledge of drug trends and emerging substances tends to be informed by data from postmortem toxicology, police seizures, urinalysis, and qualitative studies. These methods are often delayed and encumbered by limitations of selection bias and representativeness, whereas community drug checking offers faster actionable insights at the local level (11,12).

Drug checking is a voluntary process where an individual’s substance is analyzed to identify its components, including unexpected or harmful adulterants (11,15,17–19). By revealing what is actually in a substance, drug checking helps prevent accidental overdose and other negative consequences while supporting safer use decisions (13,14). U.S. overdose deaths disproportionately affect marginalized communities, particularly Black and American Indian/Alaska Native populations, making culturally attuned, community-based approaches like drug checking a way to foster trust, strengthen engagement, reduce stigma, and encourage peer-to-peer knowledge sharing in what can be isolated and underserved communities (20–22).

Community drug checking also generates critical information about the overall drug supply. In the U.S., programs range from formal public health partnerships to grassroots, peer-led initiatives, demonstrating how drug checking functions simultaneously as a public health service and a source of data to inform health strategies (21,23).

In contrast, police may employ field-ready drug testing technologies similar to those used in drug checking, but results are typically not shared with the community (11,26). While police drug seizures do not yield a robust random sample, a study comparing forensic, consumer drug checking, and poison control samples in the Netherlands found consistency in detection of novel drugs across the sources at a national level (27). No study has compared these data sources in the U.S. setting, where drug checking is nascent and fentanyl and emergent adulterants dominate the drug supply.

Using data from police department (PD) and community partner (CP) drug samples collected from eight Massachusetts communities, we compared expected and confirmed substance types (e.g., expected to be cocaine; confirmed as fentanyl), drug form (e.g., powder, pills), packaging (e.g., wax folds, baggies), and sociodemographic characteristics of the individuals whose samples were analyzed.

## MATERIALS AND METHODS

The Massachusetts Drug Supply Data Stream (MADDS) is a statewide program commenced in 2019 that provides drug checking services within CPs and also tests seized samples at PDs (13). Brandeis provides MADDS training and technical assistance. While CP staff were trained by Brandeis to conduct drug checking, Brandeis drug checking technicians performed all testing of seized samples at PDs. As of 2025, the MADDS program served 19 CPs and 8 PDs. Communities with both drug checking and PD data comprise this study’s setting, i.e., the cities of Beverly, Fall River, Gloucester, Lawrence, Lynn, New Bedford, Quincy, and Worcester. Drug samples from both sources in each community were collected and tested using standardized procedures (13). CPs submitted remnant drug material, including small amounts of powder or pills, as well as materials such as cookers or pipes containing drug residue.

Samples were either donated by PWUD to local CPs, or by PDs in compliance with an established protocol. As in our prior validation studies, and to improve PD comparability to CP samples (13,43), PD samples consisted of personal-use quantities. This included exhibits from cases with ≤ 1 g of a drug, small-scale distribution (<50 bags), or, if charged with manufacturing with intent to distribute 50 or more bags, one representative sample from such an exhibit was tested. Samples also included similar quantity exhibits from controlled buys obtained during narcotics investigations, from an overdose or other health emergency where police seized drugs, and those logged as found property. PD samples were deemed to serve no further investigative purpose and would otherwise be sent for destruction.

### Testing and analytic procedures

Samples were tested by trained staff using immunoassay test strips and Fourier-transform infrared spectrometry (FTIR) then by Gas Chromatography/Mass Spectrometry (GC/MS) at an off-site laboratory (for details see (13)). Following completion of testing, results were shared via StreetCheck, a public website that displays sample-specific results and aggregate drug supply trends (44). CPs receive access to individual, sample-level findings, whereas partnering PDs see aggregate results.

Samples included in the current analysis were collected and tested between April 1, 2018 and October 30, 2025, with laboratory results received through December 31, 2025.

### Measures

Measures included characteristics of drug samples and submitters.

#### Suspected and confirmed sample components

We provided categories for suspected samples that were reported as (a) suspected and (b) unknown at the time of sample collection. What the substance was suspected to be was ascertained from the sample’s donor at a CP or extracted from PD case reports. Confirmed sample components were the active substances found by laboratory testing. We compared the laboratory-confirmed substances and the suspected substances at the time of sample collection for powder and crack cocaine, opioids, methamphetamine, benzodiazepines, and xylazine. This variable was operationalized across each substance as (a) suspected and confirmed or (b) not suspected but confirmed. Multiple substances could be confirmed in one sample.

#### Relative ratios of lab-derived components

Test results included the relative amount of each active substance in a sample derived from testing. We first compared the amount of fentanyl in samples relative to other substances, indicated as (a) fentanyl as the primary substance, (b) fentanyl as equally present as one or more other active substances, and (c) fentanyl present in amounts less than one or more other active substances. We then identified how many samples contained both fentanyl and xylazine, and the number of samples where stimulants, including cocaine, methamphetamine, or amphetamines, were present with fentanyl. We further parsed out fentanyl-contaminated stimulants by first identifying laboratory-confirmed samples containing the stimulants of cocaine, methamphetamine, or amphetamines as the primary substance, while removing samples where packaging (e.g., cookers, cottons, and pipes) could have contained residual fentanyl from prior use.

#### Time to detect emergent substances and drug patterns of concern

For emerging substances (i.e., xylazine, medetomidine, bromazolam, nitazenes) and a concerning drug pattern (i.e., contamination of stimulants with fentanyl defined as a sample where fentanyl is detected and the primary substance identified is a stimulant and submitted as raw drug (i.e., no packaging, not residue)), we report the month and year in which each was first detected in PDs and CPs across the eight locations.

#### Drug form and packaging

At the time of collection or testing, trained technicians report the form, operationalized as powder, residue, pill, rock, or other, as well as the packaging, operationalized as baggie or fold, cookers and/or cottons, pipes and chores, other drug packaging (e.g., straws, foils, vials), multiple packaging types, and no packaging.

#### Submitter demographics, sample location, and associated adverse events

At submission, submitters could report gender, race/ethnicity, and whether the sample had an associated fatal or nonfatal overdose event. Location of the sample at time of collection was logged for PD samples only. Variables were operationalized for analysis as: man or woman; White, Black, Hispanic/Latinx, or other races/ethnicities; home or residence, public place, institution/business, and other (e.g., traffic stop); and reported fatal or nonfatal overdose.

#### Pills

Pill samples were characterized based on component characteristics and indications of counterfeit status. We considered the substances detected in pill samples, operationalized as none, one, and two or more. We operationalized counterfeit pills as (a) suspected substance aligned with laboratory-confirmed substance, (b) suspected substance was discrepant with laboratory-confirmed substance, and (c) suspected substance was reported as unknown. We also explored the primary and secondary laboratory-confirmed substances present in pill samples: benzodiazepines, fentanyl, caffeine, acetaminophen, amphetamines, prescription opioids (e.g., oxycodone, hydrocodone), stimulants (methamphetamine, cocaine), xylazine, other (e.g., local anesthetics, anti-seizure medications).

### Analysis

Descriptive and inferential statistics were computed using R (Version 4.5) on all sample and submitter characteristics reported by sample source. To test for differences in PD versus CP samples, we used Chi-square tests of independence. Fisher’s exact tests (FET) and their p-values were reported with expected cell sizes <5. For larger crosstabulations, uneven expected cell sizes, and a global test of association, Freeman–Halton extension (FET-FHE) with Monte Carlo simulation (B = 100,000) was used. We calculated the difference in days between detection of each emerging substance or concerning drug pattern in a CP site and their paired PD, using the date of testing as well as the date of acquisition. Median and interquartile range in detection times across substances were also calculated.

This study was approved by the Brandeis University and Massachusetts Department of Public Health Institutional Review Boards.

## RESULTS

During the study period, there were 2,430 unique samples submitted by both CPs and PDs in eight different Massachusetts locations for which laboratory testing was conducted; 68.1% originated from CPs and 31.9% derived from PDs.

### Compositional characteristics of drug samples

Across all samples, fentanyl and cocaine were the most common substances identified (Table 1). Overall, there were more samples derived from CPs for any given substance type than from PDs. More samples submitted through CPs contained cocaine, fentanyl, xylazine, methamphetamine and medetomidine than those derived from PDs; there were no differences by source for heroin, benzodiazepine or nitazene samples.

**Table 1.** Compositional characteristics of drug samples collected by police departments and community partners in eight Massachusetts cities.

| | Total<br>(N=2,430) | Police<br>Department<br>(N=775) | Community<br>Partner<br>(N=1,655) | $\chi^2$ or FET |
| --- | --- | --- | --- | --- |
| *Compositional characteristic* | N (%) | n (%) | n (%) |  |
| **Laboratory-confirmed drug sample components<sup>1</sup>** |  |  |  |  |
| Cocaine (Powder or Crack) | 1,042 (42.9) | 222 (28.7) | 820 (49.5) | 94.1*** |
| Fentanyl | 1,619 (66.6) | 474 (61.2) | 1,145 (69.2) | 15.3*** |
| Heroin | 115 (4.7) | 31 (4.0) | 84 (5.1) | 1.4 |
| Methamphetamine | 143 (5.9) | 35 (4.5) | 108 (6.5) | 3.8* |
| Benzodiazepines | 108 (4.4) | 41 (5.3) | 67 (4.0) | 1.9 |
| Xylazine | 569 (23.4) | 116 (15.0) | 453 (27.4) | 45.3*** |
| Medetomidine | 42 (1.7) | 5 (0.6) | 37 (2.2) | p<.01 |
| Nitazenes | 5 (0.2) | 1 (0.1) | 4 (0.2) | p=1.00 |
| **Suspected samples** |  |  |  | 241.8*** |
| *Known* | 1,887 (77.7) | 453 (58.5) | 1,434 (86.6) |  |
| *Unknown* | 543 (22.3) | 322 (41.5) | 221 (13.4) |  |
| **Suspected vs. confirmed sample components<sup>2</sup>** |  |  |  |  |
| *Cocaine (Powder & Crack) (n=1,042)* |  |  |  | 0.01 |
| Suspected and confirmed | 419 (40.2) | 90 (40.5) | 329 (40.1) |  |
| Not suspected, but confirmed | 623 (59.8) | 132 (59.5) | 491 (59.9) |  |
| *Opioids<sup>2</sup> (n=1,626)* |  |  |  | 105.9*** |
| Suspected and confirmed | 1,186 (72.9) | 264 (55.3) | 922 (80.2) |  |
| Not suspected, but confirmed | 440 (27.1) | 213 (44.7) | 227 (19.8) |  |
| *Methamphetamine (n=143)* |  |  |  | 6.9** |
| Suspected and confirmed | 51 (35.7) | 6 (17.1) | 45 (41.7) |  |
| Not suspected, but confirmed | 92 (64.3) | 29 (82.9) | 63 (58.3) |  |
| *Benzodiazepines (n=108)* |  |  |  | 0.6 |
| Suspected and confirmed | 45 (41.7) | 19 (46.3) | 26 (38.8) |  |
| Not suspected, but confirmed | 63 (58.3) | 22 (53.7) | 41 (61.2) |  |
| *Xylazine (n=569)* |  |  |  | NA |
| Suspected and confirmed | 0 (0) | 0 (0.0) | 0 (0.0) |  |
| Not suspected, but confirmed | 569 (100.0) | 116 (100.0) | 453 (100.0) |  |
| **Relative ratios** |  |  |  |  |
| *Fentanyl vs. all other cuts (N=1,523)* |  |  |  | 16.8*** |
| Primary substance | 1,019 (66.9) | 325 (74.2) | 694 (64.0) |  |
| Equal to primary substance | 87 (5.7) | 25 (5.7) | 62 (5.7) |  |
| Less than primary substance | 417 (27.4) | 88 (20.1) | 329 (30.3) |  |
| **Both fentanyl and xylazine present** | 559 (23.0) | 113 (14.6) | 446 (26.9) | 45.6*** |
| **Both fentanyl and stimulants present** | 569 (23.4) | 70 (9.0) | 499 (30.2) | 131.3*** |
| **Fentanyl-contaminated stimulants (n=569)** | 31 (5.4) | 9 (1.6) | 22 (3.9) | p=0.85 |

**Notes:** FET = Fisher's Exact Test; p-values were reported when Fisher's Exact tests were used (i.e., medetomidine and nitazenes).
\*p<.05, \*\*p<.01, \*\*\*p<.001
<sup>1</sup>Categories are not mutually exclusive; each drug sample may be represented in more than one category. <sup>2</sup>Opioids suspected in these samples include fentanyl, heroin, dope, and morphine and were then compared to samples that were confirmed to contain fentanyl and heroin.

When comparing the suspected substances within samples reported by PDs and CPs relative to the laboratory-confirmed results, samples containing opioids and methamphetamine submitted through CPs were more likely than PD samples to be suspected and laboratory-confirmed to contain these substances than confirmed but not suspected (Table 1). Regardless of sample source (i.e., PD vs. CP), few suspected xylazine and methamphetamine in drug samples, despite subsequent laboratory-confirmed presence of these substances. Overall, opioid samples had the most alignment between suspected and confirmed results. Irrespective of sample components, PD samples were significantly more likely than community samples to be of “unknown” content.

Fentanyl, relative to other substances, was confirmed as the primary substance in more PD than CP samples. Regardless of source, in samples where both fentanyl and other active substances were present, fentanyl more commonly appeared in greater proportions than equal to or lower proportions than other active substances (Table 1). Samples where both fentanyl and xylazine were detected were more common among samples from CPs than samples submitted by PDs. Both stimulants and fentanyl were detected together in approximately a quarter of all samples (23.4%) and were more commonly present together among community rather than PD samples. Regardless of source, of all samples containing stimulants and fentanyl, 5.4% were identified as samples with a stimulant as its primary substance that also contained fentanyl.

### Emergent Substances and Drug Patterns of Concern

Figure 1 shows that, across the study sites, four emergent substances and one harmful, unexpected pattern of drugs were detected a median of 249 days earlier [IQR 52.5, 732] in CP samples compared to PD seizure samples based on their test date. If, instead, the samples had been tested at the time of acquisition, detection of the substances or pattern would have been 199 [IQR 84, 632] days earlier via CP samples compared to PD seizure samples. The time windows were smaller and more closely aligned for CPs and PDs when considering detection of the presence of fentanyl in stimulants (Figure 1).

**Figure 1.**
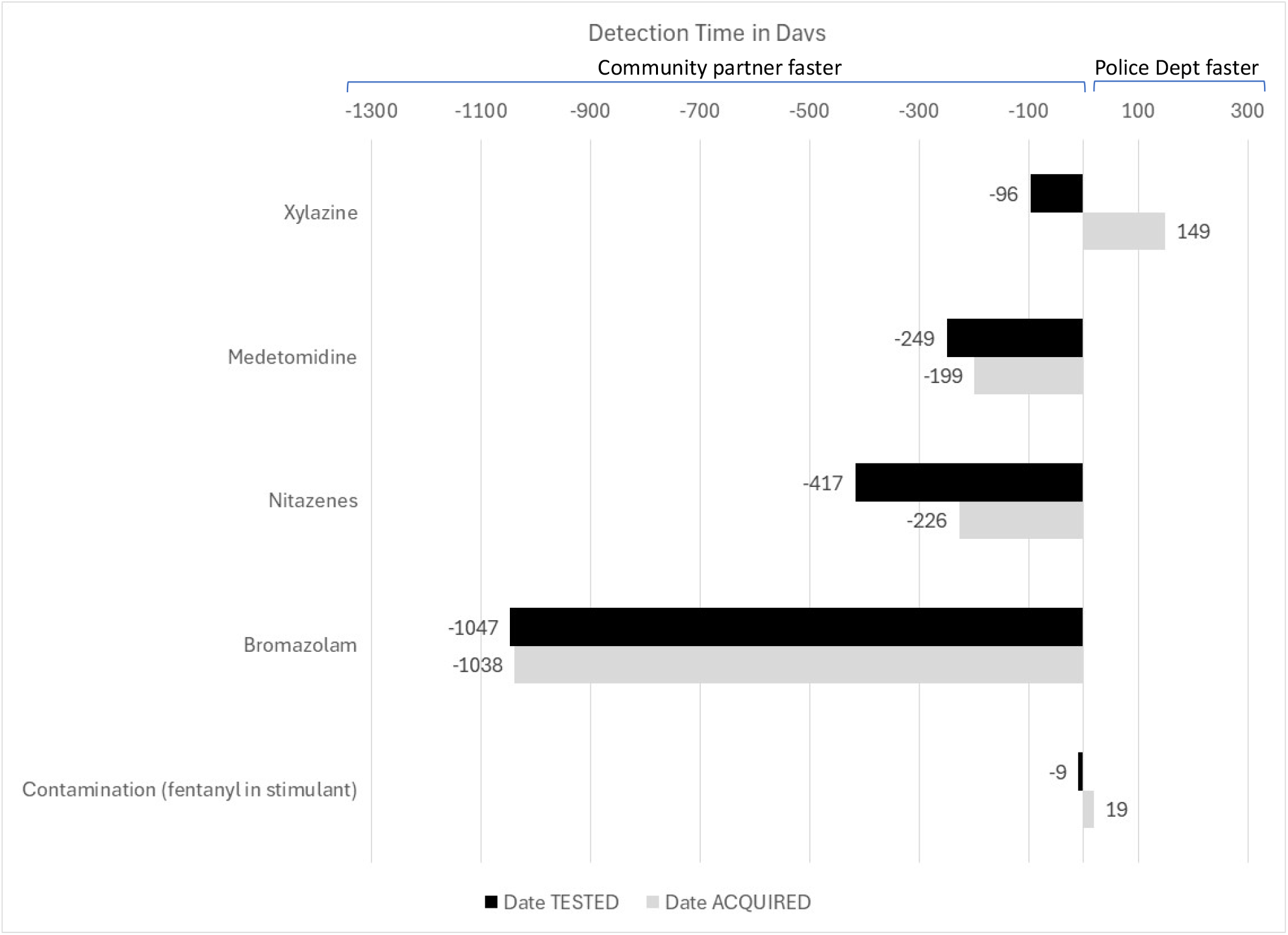
Differences in time to detection by substance: Paired community partner (-) and police department (+) samples *For the first detection of bromazolam (a community partner site on 4/2021), there was no paired police department sample tested where bromazolam was detected. However, a study site nearby also detected bromazolam soon afterward on 6/2021 and their police department samples were used for the bromazolam calculations.

### Drug form and packaging

The form in which samples were submitted for drug checking significantly differed by source. Most samples provided by PDs were powder, whereas most samples submitted through CPs were residues. Pills and rock were more common in PD samples than those submitted through CPs (Table 2). The packaging type also significantly differed by sample source. Community samples were submitted in a wider array of drug packaging types relative to PD samples, with representation in baggies and folds, cookers and/or cottons, pipes and chores, and without packaging. Samples derived from paraphernalia were rare among PD samples; most were collected or submitted within baggies or folds.

**Table 2.**
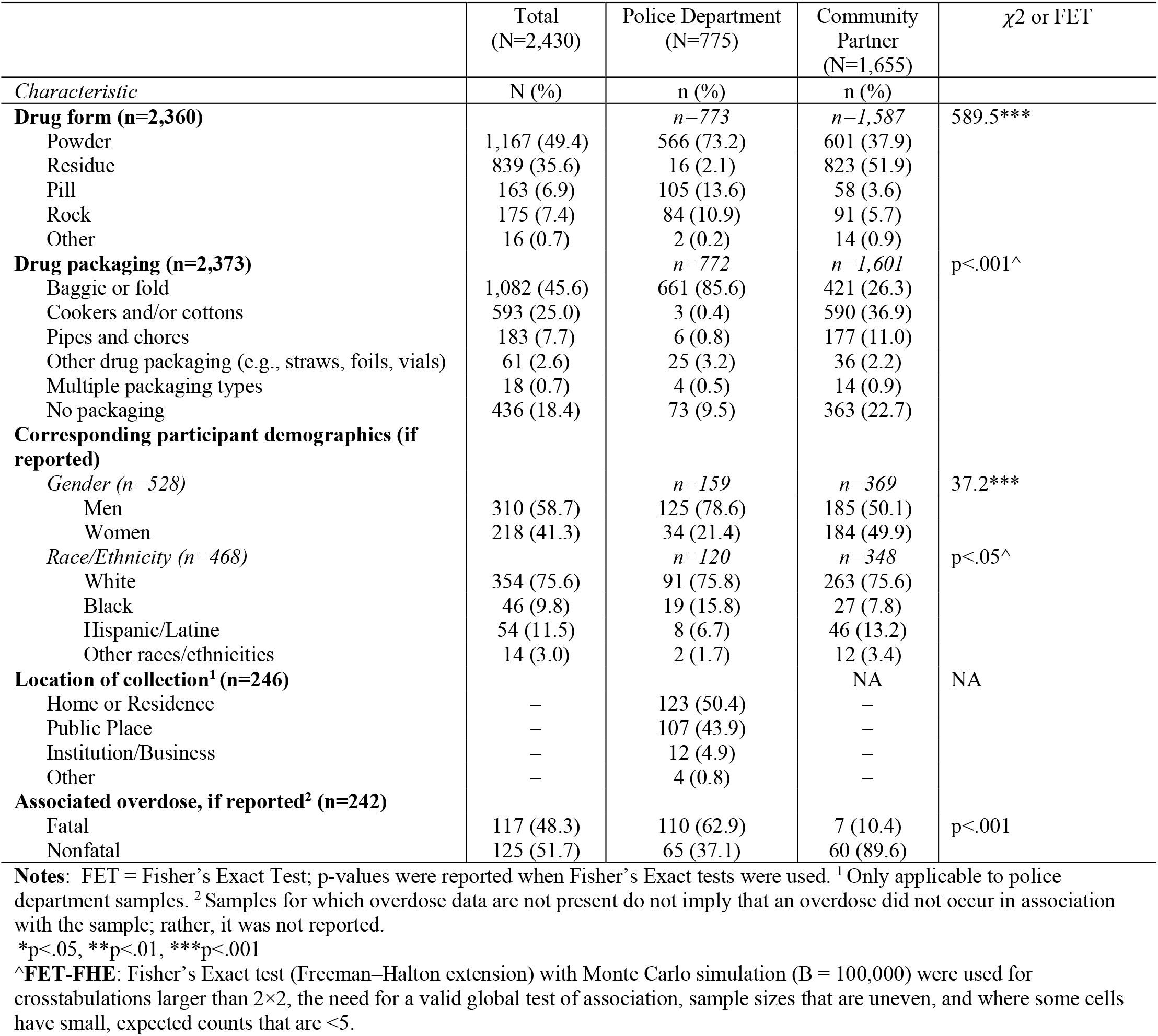
Other characteristics of drug samples collected by police departments and community partners in eight Massachusetts cities.

### Submitter demographics and sample location

The gender of those submitting drug samples was reported in 21.7% of samples and race/ethnicity was reported in 19.3% of samples (Table 2). Samples from PDs were significantly more likely to come from men than CP samples. While White individuals were represented similarly between PD and community samples, more Black individuals were implicated with drug samples originating from PDs than CP samples, and more Hispanic/Latine individuals were associated with CP than PD samples.

PD samples were seized equally from private residences and public places, followed by institutions/businesses and other locations such as traffic stops.

### Overdose

Associated fatal overdose of a submitted drug sample was reported in 4.8% of samples, and 5.1% of samples were associated with non-fatal overdose. The proportion of samples associated with any type of overdose event was higher for PD than community samples (Table 2). Samples associated with non-fatal overdose were more common among community samples than PD samples, whereas samples associated with fatal overdose were more likely sourced from PDs than CPs.

### Pills

Approximately 7% of samples overall were pills. Proportionally, pills were more frequently found in PD than CP samples (Table 2). Community pill samples were equally aligned and discrepant in suspected relative to confirmed pill identity, whereas pill samples believed to be counterfeit from PDs were reported as less discrepant but were more likely to be submitted without a suspected substance readily identified (i.e., unknown pill type) compared to CP pill samples (Table 3).

**Table 3.** Characteristics of pill samples collected by police departments and community partners in eight Massachusetts cities (N=163)

|  | Total<br>(N=163) | Police Department<br>(N=105) | Community<br>Partner<br>(N=58) | FET-FHE |
| --- | --- | --- | --- | --- |
| *Characteristic* | N (%) | n (%) | n (%) |  |
| **Number of active substances present in pill (n=163)** |  |  |  | p=0.61 |
| None | 6 (3.7) | 3 (2.9) | 3 (5.2) |  |
| One | 90 (55.2) | 60 (57.1) | 30 (51.7) |  |
| Two or more | 67 (41.1) | 42 (40.0) | 25 (43.1) |  |
| **Counterfeit pills (n=159)<sup>1</sup>** |  |  |  | p<.001 |
| Suspected aligned w/ confirmed | 68 (42.8) | 38 (37.2) | 30 (52.6) |  |
| Suspected and confirmed are discrepant | 46 (28.9) | 21 (20.6) | 25 (43.8) |  |
| Suspected was reported as unknown | 45 (28.3) | 43 (42.2) | 2 (3.6) |  |
| **Primary confirmed substance (n=149)** |  |  |  | p<.05 |
| Benzodiazepines | 53 (35.6) | 34 (35.8) | 19 (35.2) |  |
| Fentanyl | 16 (10.7) | 11 (11.6) | 5 (9.2) |  |
| Caffeine | 12 (8.1) | 11 (11.6) | 1 (1.9) |  |
| Acetaminophen | 17 (11.4) | 7 (7.4) | 10 (18.5) |  |
| Amphetamines (e.g., Adderall) | 11 (7.4) | 9 (9.5) | 2 (3.7) |  |
| Rx opioids (e.g., oxycodone, hydrocodone) | 10 (6.7) | 7 (7.4) | 3 (5.5) |  |
| Stimulants (methamphetamine or cocaine)<sup>3</sup> | 5 (3.3) | 1 (1.0) | 4 (7.4) |  |
| Xylazine | 4 (2.7) | 3 (3.1) | 1 (1.9) |  |
| Other (e.g., anesthetics, anti-seizure)<sup>2</sup> | 18 (12.1) | 12 (12.6) | 6 (11.1) |  |
| Multiple primary confirmed substances | 3 (2.0) | 0 (0.0) | 3 (5.6) |  |
| **Secondary confirmed substance (n=60)** |  |  |  | p<.05 |
| Benzodiazepines | 1 (1.7) | 0 (0.0) | 1 (4.8) |  |
| Fentanyl | 13 (21.7) | 5 (12.8) | 8 (38.1) |  |
| Caffeine | 3 (5.0) | 1 (2.6) | 2 (9.5) |  |
| Rx opioids | 2 (3.3) | 1 (2.6) | 1 (4.8) |  |
| Stimulants (methamphetamine or cocaine)<sup>3</sup> | 17 (28.3) | 12 (30.8) | 5 (23.8) |  |
| Xylazine | 1 (1.7) | 1 (2.6) | 0 (0.0) |  |
| Other (e.g., anesthetics, anti-seizure)<sup>4</sup> | 15 (25.0) | 14 (35.9) | 1 (4.8) |  |
| Multiple secondary confirmed substances | 8 (13.3) | 5 (12.8) | 3 (14.3) |  |

**Notes:** FET-FHE = Fisher's Exact test (Freeman-Halton extension) with Monte Carlo simulation (B = 100,000) were used for crosstabulations larger than 2×2, the need for a valid global test of association, sample sizes that are uneven, and where some cells have small, expected counts that are <5. p-values were reported when Fisher's Exact tests were used.
<sup>1</sup>Four pill samples were missing any information about what substances were suspected. <sup>2</sup>Other primary substances include anti-histamines, phenacetin, muscle relaxants, anti-depressants, hormones, and more. <sup>3</sup>The vast majority of stimulant secondary substances were comprised of methamphetamine. <sup>4</sup>Other secondary substances include: Nitazenes, tramadol, dietary supplements, and more.

Regardless of source, over half (55.2%) of pill samples consisted of one active substance while 41.1% contained two or more active substances (Table 3). The primary confirmed substance was most often a benzodiazepine, followed by fentanyl, albeit a diverse array of other substances— caffeine, acetaminophen, amphetamines, prescription pain medication, xylazine and more—were identified as primary substances across both PD and CP samples. For pills containing more than one substance, fentanyl was the second most common confirmed substance, followed by stimulants (e.g., methamphetamine, cocaine).

## DISCUSSION

As challenges of the synthetic drug crisis continue to evolve in the U.S., the need for understanding the composition of the drug supply is ever greater. This analysis found that PD and CP samples provide unique, complementary information about the drug supply. The information is useful for monitoring trends, with implications for emerging substance detection and characterizing evolving definitions of drug-specific harm.

Different from prior state-based database comparisons, our study derives drug samples from different sources and holds constant the testing procedures and analytic methods, suggesting that the detected results may be attributable to true differences in the sources themselves rather than to variability in the process of testing or other external factors. Moreover, all prior studies comparing forensic seized drug results to poison control or clinical drug data relied upon state-generated datasets of seized drugs, which are often of unknown origin and lack incident, demographic or health indicator variables (27,46). Uniquely, the extractions of sample, event, and demographic characteristics from the current undertaking are rich additions to the analysis of drug supply data and speak to the needs for both better and more consistent incident reporting as well as data systems that systematically extract them for monitoring.

We found that samples from PDs and CPs significantly differed in ways that impact inferences and generalizability about the drug supply. CP sample submissions aligned more often in suspected and tested content than PD samples, tended to identify, catalogue and contain emergent adulterants, and were less frequently associated with fatal and nonfatal overdose. PD samples tended to contain more raw drug in both powder and pill form, and accessed samples with fentanyl as the primary substance. Hence, taken at face value, PD samples may portray a more lethal, less complex drug supply than experienced by PWUD in their community. Differences by source in drugs suspected could be the result of distinct collection and documentation methods, as CPs interact with PWUD seeking to understand the contents of their supply (21,23,47), while PDs typically collect samples through controlled buys, search warrant seizures, found property, and overdose events, with limited information obtained from the people who possessed the drugs (12). Such encounters are more likely to yield drugs in whole form rather than as residue, and more conservative estimates of what the samples may be composed of. Additionally, PDs may employ different reporting methods; standardized protocols would improve data quality and completeness.

There was a meaningful and concerning delay in detection of emergent substances when comparing CP and PD seized samples of drugs. Access to PD-based samples may be delayed due to administrative protocols, investigative and prosecutorial timelines, and staffing or resource constraints. For instance, a sample containing xylazine collected in January 2020, when the substance was emergent, was not tested until September 2020. Similarly, an active investigation in one site ultimately led to many arrests for suspected counterfeit pill manufacturing, which may have contributed to the dearth of PD samples available for testing that contained bromazolam. Had drug supply monitoring relied exclusively on PD samples, one may have concluded erroneously that the regions were unaffected by xylazine or bromazolam. These observations illustrate inconsistencies and biases that may be addressed by centering community drug checking samples as a critical community early warning source. Moreover, studies report the effectiveness of community drug checking programs in helping PWUD make informed decisions about safer use (48–50). Our analysis suggests that CP programs are well positioned to detect adulterants and novel substances and provide real-time public health guidance directly to PWUD (51–53).

For a more comprehensive understanding of the drug supply, however, examining samples from PDs may provide data not otherwise captured through community-based collection alone. Sample presentation differences showed that most samples tested at PDs were identified in powder, pill, or rock form and within baggies, whereas CPs more often received samples in residue form. These differences likely reflect the distinct points of collection within the drug supply chain (54), as PDs often intercept drugs prepared for distribution, while CPs receive samples directly before or after consumption. PD sample testing uniquely indicates, for instance, that 5.4% of powder stimulant samples contain fentanyl, suggesting a low, local contamination level that may be difficult to derive from CPs testing mostly remnant samples. Additionally, pills were more commonly tested at PDs and rarely submitted for testing at CPs, making PD samples informative for assessing trends in counterfeiting or, more generally, uncovering supply risks for people who do not frequent syringe service or overdose prevention program-based drug checking services.

Our analysis also identified sociodemographic differences in samples tested at CP and PD drug checking sites that have methodological implications when considering bias and representativeness. Consistent with research highlighting disproportionate representation of people of color throughout the U.S. criminal-legal system, we found that samples collected by PDs were more commonly associated with Black individuals. Similarly, PDs obtained proportionally more samples from men than women compared to CPs (55–58). However, racial, ethnic, and gender disparities in syringe service and overdose prevention program participation have been documented (59,60). As such, our findings reflect that CPs may be missing important demographics in their drug checking initiatives (61,62), and should consider strategies that reach populations who are less likely to access traditional syringe service and overdose prevention services (63).

Given limited resources, one may question whether to expand community drug checking or invest in monitoring of PD-seized samples to gather knowledge about the drug supply. While community drug checking excels in more rapid detection of new and emerging substances, our analysis suggests both sources contribute meaningfully to improving a community’s ability to characterize what, where, and who is affected by evolving drug supplies. Systematic testing of samples could facilitate a comprehensive, multi-pronged approach that encompasses both sources. Additionally, further research is needed to determine if and how PD and CP programs can effectively share information about the drug supply. If done compassionately, PDs—in complementing the critical efforts of CPs—could convey drug supply information to help PWUD make safer, more informed decisions about substance use.

### Limitations

Findings may not be generalizable given convenience sampling from the eight cities in which both PD and CP samples were available in our setting. Nonetheless, given the present barriers to accessing drug checking services (e.g., legal ramifications of drug possession and distribution), well-powered convenience samples can play an important role in shaping practice and policy about the nation’s unpredictable illicit drug supply. Significant differences by source characteristics may arise from the reasons they were collected (e.g., curiosity among PWUD about substance composition vs. the goal of interrupting the distribution and consumption of substances in a manner comporting with legal requirements) and may influence detection. Additionally, reported sociodemographics of sample submitters was low. Future drug checking efforts could adopt systematic, confidential methods of collecting these data.

## CONCLUSION

Local drug supply data from CP drug checking programs and PD seizures generate complementary information about drug trends. Differences in these sources’ sampling and characteristics may introduce bias when presenting trends in isolation, but each offers unique insights into drug epidemiology, for cataloging emerging drugs, and in preparing health system responses.

## Data Availability

All data produced in the present study are available upon reasonable request to the authors and with permission from the Massachusetts Department of Public Health

http://www.streetcheck.org

